# The influence of calcific aortic stenosis on atrioventricular conduction: a prospective observational study and Mendelian randomization

**DOI:** 10.1101/2024.05.08.24307094

**Authors:** Karan Rao, Jonathan L Ciofani, Daniel Han, Milad Nazarzadeh, Kazem Rahimi, Alexandra Baer, Natasha Saad, Princess Neila Litkouhi, Usaid Allahwala, Peter Hansen, Ravinay Bhindi

**Author notes:** Karan Rao and Jonathan L Ciofani are joint first authors. Correspondence to: Professor Ravinay Bhindi, Department of Cardiology, Royal North Shore Hospital, Reserve Road, St Leonards 2065 Australia.

## Abstract

**Background:** Aortic stenosis (AS) may prolong atrioventricular (AV) conduction secondary to calcium infiltration of the atrioventricular-His-pathway, which is hypothesized to contribute to bradyarrhythmia and sudden cardiac death in this cohort. This study investigates the correlation between calcific AS and impaired AV conduction.

**Methods:** We performed an observational study and Mendelian randomization (MR). The observational study was a sub-analysis on AS patients undergoing transcatheter aortic valve implantation2021-2023). Regression analyses were used to assess the association between aortic valve calcium volumes with electrophysiology study derived markers of AV conduction. The primary MR analyses used European-ancestry genome-wide association studies for AS (653,867 participants,13,765 cases) and PR interval (271,570 participants). The exposure outcome was the genetic liability for AS, whilst the outcome was prolonged PR interval or AV block.

**Results:** The mean age in the observational study (n=196) was 81.8 years, 64.8% were men, and 46(23.6%) had first-degree heart block. There was no significant association between total aortic valve complex calcium volume and first-degree heart block, or with PR, AH or HV intervals. On MR, no association was found between genetic liability for AS and PR interval (beta -0.41, 95%CI -1.33-0.51, p=0.38). Additional analyses evaluating genetic liability for AS and AV block suggested a significant association on IVW analysis (OR 1.15, 95%CI 1.02-1.30, p=0.02), but this was not supported by sensitivity analyses and thus interpreted with caution.

**Conclusions:** This study found no meaningful association between aortic valve calcification and PR, AH or HV interval, challenging the hypothesis that calcific AS impairs atrioventricular conduction.

## Introduction

Aortic stenosis (AS) is the most common valve pathology affecting 12.4% of the population over the age of 75 years^1^. It is typically degenerative, involving fibro-calcific remodelling of the aortic valve complex resulting in leaflet restriction. When AS is severe and untreated, it can have a mortality up to 50% at 5 years, with an incidence of 9.2% for unexplained sudden cardiac death^2^. Reduced cardiac output, progressive myocardial fibrosis and ischaemia, as well as abnormal atrioventricular (AV) conduction^3^ have been proposed as contributors to mortality.

Abnormal atrioventricular conduction has been hypothesized to be driven by left ventricular hypertrophy or calcium extension into the interventricular septum. The AV-His-Purkinje pathway lies near to the aortic valve complex (AVC). Specifically, the His bundle traverses and bifurcates at the membranous interventricular septum which is located within the interleaflet triangle between the non and right coronary (aortic) cusps. In cases of severe AS, it is hypothesized that valvular calcification could extend into the neighbouring conduction pathway, potentially disrupting AV nodal conduction. This association has been explored previously in small-scale historical studies, where a correlation between calcific AS and delays in PR interval, His bundle, and interventricular conduction has been observed^3–5^. However, there is a paucity of modern data involving detailed assessment of calcium volume distribution in the aortic valve apparatus and its as association with atrioventricular conduction.

This study aims to investigate the association of calcific AS with AV conduction through an observational study and Mendelian randomization (MR) method. The observational study aims to study patients with known AS and correlate the imaging derived volume and distribution of aortic valve calcium with invasively measured markers of AV conduction.

However, considering the unavailability of an appropriate propensity-matched control arm, the observational study is prone to reverse causation and confounding, which may partially be overcome through MR. MR allows us to investigate the potentially causal relationship between the exposure variable of AS, and the outcome variable of atrioventricular conduction delay. MR relies on the premise that genetic polymorphisms contribute to an individual’s phenotype and the random inheritance of these phenotype-determining genetic polymorphisms is analogous to assignment to a treatment group in a randomised control trial^6^, thus aiming to mitigate reverse causation and confounding. This approach has previously been used to demonstrate potentially causal associations between risk factors and cardiovascular diseases including coronary disease and AS^7,8^, as well as between common cardiovascular risk factors and AV block^9^.

## Methods

### Prospective Observational Study

#### Study Design

This study included 196 patients with symptomatic severe AS from November 2021 until October 2023, and performed as a sub-analysis on data which was prospectively collected from an active clinical study on patients awaiting transcatheter aortic valve implantation (TAVI)^10^ (CONDUCT TAVI: ACTRN1261001700820) at a large tertiary referral hospital in Sydney, Australia. Those with a pre-existing permanent pacemaker, or with prior aortic valve surgery were excluded. The study was approved by Northern Sydney Local Health District Ethics Committee.

#### Computed Tomography (CT) Analysis

All patients underwent multi-detector CT angiography. Image acquisition and sequences were per standard pre-TAVI protocols, and not mandated by the study. Images were analysed offline, using 3Mensio medical imaging software (Pie Medical Imaging, Netherlands). Volumetric calcium analyses of the aortic valve complex using pre-specified variables was performed. The primary analysis was performed on the aortic valve complex (AVC), which was defined as the combination of the basal leaflets (first 10mm from annular plane to aortic root), and the upper LVOT (from annular plane to 5mm into LVOT) (Figure 1). Multiple secondary analyses were performed on differential calcium volumes split by location (LVOT) as well as by aortic cusps (non-coronary cusp, left coronary cusp and right coronary cusp) (Figure 1). The Hounsfield (HU) unit threshold for calcium detection was manually adjusted for each scan to ensure accurate and comprehensive capture of calcification.

**Figure 1:**
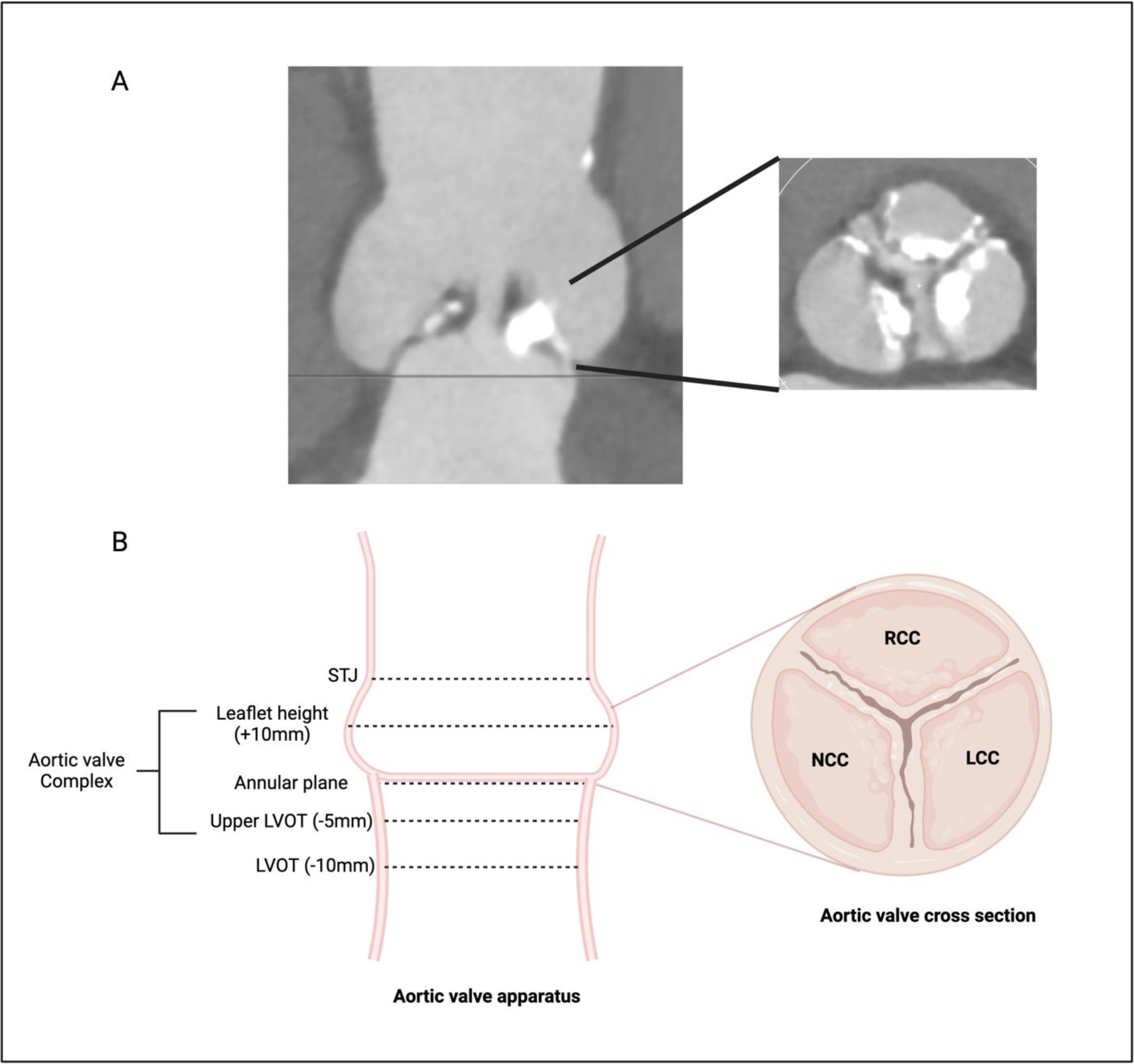
CT based volumetric analysis of the aortic valve apparatus. Part A demonstrates a patient’s longitudinal stretched aorta vessel view (left) and the cross-section (right). Part B is a labelled schematic diagram of the same. The aortic valve complex was defined as upper LVOT (-5mm from aortic annulus) to leaflet height (+10mm from annular plane). LVOT defined as 10mm below the annular plane. RCC: right coronary cusp; NCC: non coronary cusp; LCC: left coronary cusp.

#### 12 Lead ECG and Electrophysiology Study

Atrioventricular conduction was assessed using non-invasive 12-lead ECG, as well as targeted invasive electrophysiology testing. ECG derived PR and QRS intervals were recorded, along with the overall rhythm and presence of interventricular conduction disease.

Targeted electrophysiology studies (EPS) were performed immediately prior to the planned TAVI using a mobile electrophysiology system (BIOTRONIK, Berlin, Germany). A standard quad-polar non-deflectable electrophysiology catheter (BIOTRONIK, Berlin, Germany) was inserted using ultra-sound guided transfemoral venous access. The Atrial-His (AH) and His-Ventricular (HV) signals were documented by positioning the catheter in the lower right atrium (RA), and the His signal was identified as the intervening spike between the atrial and ventricular signal. The AH interval was recorded from the onset of the atrial signal to the onset of the His bundle signal on the His catheter recording. The HV interval was recorded from the onset of the His signal on the His catheter, to the earliest ventricular (QRS) signal on the concurrent 12 lead ECG. The intervals were logged once reproducible (+/-2ms over at least 3 beats). Notably, as these measurements were recorded prior to the TAVI procedure, all patients were under conscious sedation.

#### Statistical analysis

Statistical analyses were performed using SPSS Version 29 (IBM, United States). The primary independent variable was total AVC calcium volume (mm^3^) whilst secondary variables included pre-defined calcium parameters divided by location within the AVC (non-coronary cusp, right coronary cusp, left coronary cusp and left ventricular outflow tract). The association between predictors and categorical outcome variables was assessed via logistic regression; whilst association with continuous outcome variables was assessed via multiple linear regression with adjustment for perceived confounders. A significant (p value) threshold of 0.05 was used.

### Mendelian randomisation

#### Study Design and Data Sources

The present study uses a two-sample Mendelian randomisation (MR) approach. For the main analysis, the exposure variable was genetic liability for aortic stenosis and the outcome was PR interval. Both exposure and outcome data were sourced from European ancestry populations since population homogeneity is a requirement for the samples used in a two-sample MR analysis. Three further core assumptions of MR are that the genetic variants should be: (i) strongly associated with the exposure; (ii) exclusively associated with the outcome via the exposure; and (iii) independent of potential confounders. To address the first criterion, uncorrelated genetic variants were selected that were significantly associated with the exposure at p < 5 x 10^-8^; and to account for the latter two criteria, sensitivity analyses were performed as described below.

Data for the present study are publicly available. Ethical approval and consent were obtained by the original studies. The exposure AS dataset was extracted from a genome wide association study (GWAS) meta-analysis of 10 cohorts which included 653,867 European participants (13,765 AS cases)^11^. As a sensitivity analysis, MR calculations were performed using a restricted subset of five SNPs that were also shown to be associated with aortic valve calcification (p<0.05) based on computed tomography assessment of 6,942 European participants^11^. The outcome PR interval data was from a GWAS meta-analysis of 40 cohorts including 271,570 European ancestry participants^12^. As a positive control, outcome data for heart failure was also extracted from a European GWAS meta-analysis of 486,160 participants overall, including 14,262 cases^13^. The present study was then extended by analysing the relationship between genetically predicted liability for AS and AV block, as determined by International Classification of Diseases codes in the FinnGen cohort (5,536 cases and 286,109 controls)^14^. Detailed case definitions for each study are available from the original articles (Supplementary Table 7).

Single-nucleotide polymorphisms (SNPs) that were associated with the exposure variable at p < 5 x 10^-8^ were extracted. Linkage disequilibrium clumping was performed (r^2^ < 0.001, 10 Mb distance cut-off) and the variants with the smallest p-values were selected for further analysis. F-statistics were calculated and only SNPs with values > 10 were retained for analysis.

#### Mendelian Randomisation Statistical Analyses

The primary MR analysis was an inverse variance weighted (IVW) random-effects meta-analysis of the Wald ratios for each genetic variant. Results are expressed as a beta value for genetically predicted change in PR interval duration (ms) per unit increase in log odds of genetic liability to AS. To evaluate for violation of the main MR assumptions, we conducted sensitivity analyses including weighted-median, weighted-mode, MR-Egger, MR-PRESSO and outlier exclusion by Cook’s. Leave-one-out analyses were also performed for potential outliers. Since each method has different assumptions, concordance between methods provides confidence in the conclusion. Weighted-median assumes at least half the instrumental variables are valid. Weighted-mode assumes the most common causal effect is consistent with the true effect. MR-Egger uses the Instrument Strength Independent of Direct Effect (InSIDE) assumption, requiring that the strength of pleiotropic effects from the genetic variants to the outcome are independent of the association strength between variants and exposure. The average pleiotropic effect of the genetic variants is estimated by the MR-Egger intercept. MR-PRESSO revises the estimate after outlier removal^15^. An alternative method of outlier identification was by Cook’s distance, with subsequent IVW analysis following outlier exclusion. Cochran’s Q-statistic tests for genetic variant heterogeneity. As a further sensitivity analysis, the primary and aforementioned sensitivity MR analyses were performed using a more liberal p-value cut-off for SNP inclusion of p < 5 x 10^-6^.

Statistical analyses were performed using R version 1.4.1106 with the TwoSampleMR package.

## Results

### Cross-sectional observational cohort study

From November 2021 until October 2023, 196 patients with severe AS were included in the study. The overall demographics are displayed in Table 1. The mean age was 81.8 years (+/-6.3), and 64.8% of participants were men. The median STS-PROM mortality risk score (%) was 2.78 (IQR 2.99) corresponding to low surgical risk. There was a history of atrial fibrillation in 61 (31.1%) patients, whilst 72 (36.7%) were on a beta blocker, and 12 (6.1%) were on a non-dihydropyridine calcium channel blocker at the time of ECG and EPS. The median of the mean aortic valve gradient was 42.0mmHg (IQR 15.0), whilst the median echocardiography (volume time integral) derived aortic valve area was 0.80 cm^2^ (IQR 0.29).

**Table 1:** Cohort demographics, echocardiographic, ECG and electrophysiology data BMI: body mass index; STS: Society of Thoracic Surgeons; HR: heart rate; SD: standard deviation; AV: atrioventricular; RBBB: right bundle branch block; LBBB: left bundle branch block; LAFB: left anterior fascicular block; LPFB: left posterior fascicular block.

| **Demographics** | **n = 196** |
| --- | --- |
| Age, years (mean, SD) | 81.8 (6.3) |
| Gender (Male) (n, %) | 127 (64.8) |
| BMI (kg/m<sup>2</sup>) (mean, SD) | 27.58 (5.3) |
| STS Risk Score (%) (median, IQR) | 2.78 (2.99) |
| EuroSCORE II (%) (median, IQR) | 2.33 (1.89) |
| Hypertension (n, %) | 159 (81.1) |
| Hypercholesterolaemia (n, %) | 117 (59.7) |
| Diabetes (n, %) | 55 (28.1) |
| Coronary Artery Disease (n, %) | 69 (35.2) |
| Coronary artery bypass grafting (n, %) | 20 (10.2) |
| Peripheral Arterial Disease (n, %) | 11 (5.6) |
| Previous Stroke (n, %) | 17 (8.7) |
| Chronic obstructive pulmonary disease (n, %) | 21 (10.7) |
| Chronic Kidney Disease (n, %) | 55 (28.1) |
| Any prior atrial fibrillation / flutter (n, %) | 61 (31.1) |
| Beta blocker (n, %) | 72 (36.7) |
| Non-dihydropyridine calcium channel blocker (n, %) | 12 (6.1) |
| Mean aortic valve gradient, mmHg (median, IQR) | 42.0 (15.0) |
| Aortic valve area, cm<sup>2</sup> (median, IQR) | 0.80 (0.29) |
| **Cohort ECG and electrophysiology results** |  |
| HR (bpm) (mean, SD) | 72.0 (13) |
| Presenting Rhythm (n, %) |  |
| Sinus rhythm | 113 (57.9) |
| Atrial fibrillation or flutter | 36 (18.5) |
| 1st degree AV block | 46 (23.6) |
| 2<sup>nd</sup> or 3<sup>rd</sup> degree AV block | 0 (0) |
| PR Interval (ms) (mean, SD) | 183.5 (33.1) |
| AH Interval (ms) (mean, SD) | 109.7 (34.8) |
| HV Interval (ms) (mean, SD) | 58.6 (10.9) |
| QRS Interval (ms) (mean, SD) | 100.2 (26.3) |
| Interventricular conduction delay (n, %) |  |
| None | 132 (67.7) |
| RBBB | 30 (15.3) |
| Bifascicular block + RBBB | 10 (5.1) |
| LBBB | 12 (6.1) |
| LAFB | 17 (8.7) |
| LPFB | 1 (0.5) |
| Incomplete RBBB | 1 (0.5) |
| Incomplete LBBB | 2 (1.0) |

Table 1 also specifies the ECG and EPS details of the cohort. On 12 Lead ECG analysis, 113 (57.9%) were in sinus rhythm, whilst 36 (18.5%) were in atrial fibrillation or flutter. 46 (23.6%) patients were found to be in first degree atrioventricular block, defined as a PR interval greater than 200 milliseconds (ms), whilst no patients had a *high-grade* atrioventricular block (HGAVB), defined as Mobitz II or complete heart block. The mean PR interval was 183.5ms (±33.1), and mean QRS interval was 100.2ms (±26.3). Right bundle branch block was present in 30 (15.3%) patients, whilst left bundle branch block was present in 12 (6.1%) patients.

When comparing patients with sinus rhythm to those with first degree AV block (Table 2), the age (81.2±6.9 vs. 83.0±4.9 years, respectively p = 0.11) and proportion of males (59.6 vs. 67.4%, respectively p = 0.36) were similar across both groups of patients. On echocardiography, the aortic valve mean gradient was lower in patients with first degree AV block (41.13 vs 48.07mmHg respectively, OR 0.96 [0.93-0.99], p = 0.01). There was no difference when comparing other echocardiographic characteristics including septal thickness, aortic valve area and left ventricular ejection fraction (LVEF). There was no difference between groups when comparing beta blocker or non-dihydropyridine calcium channel blocker usage (Table 2). Similarly, there was no difference between groups when comparing pre-defined CT derived calcium variables.

**Table 2:**
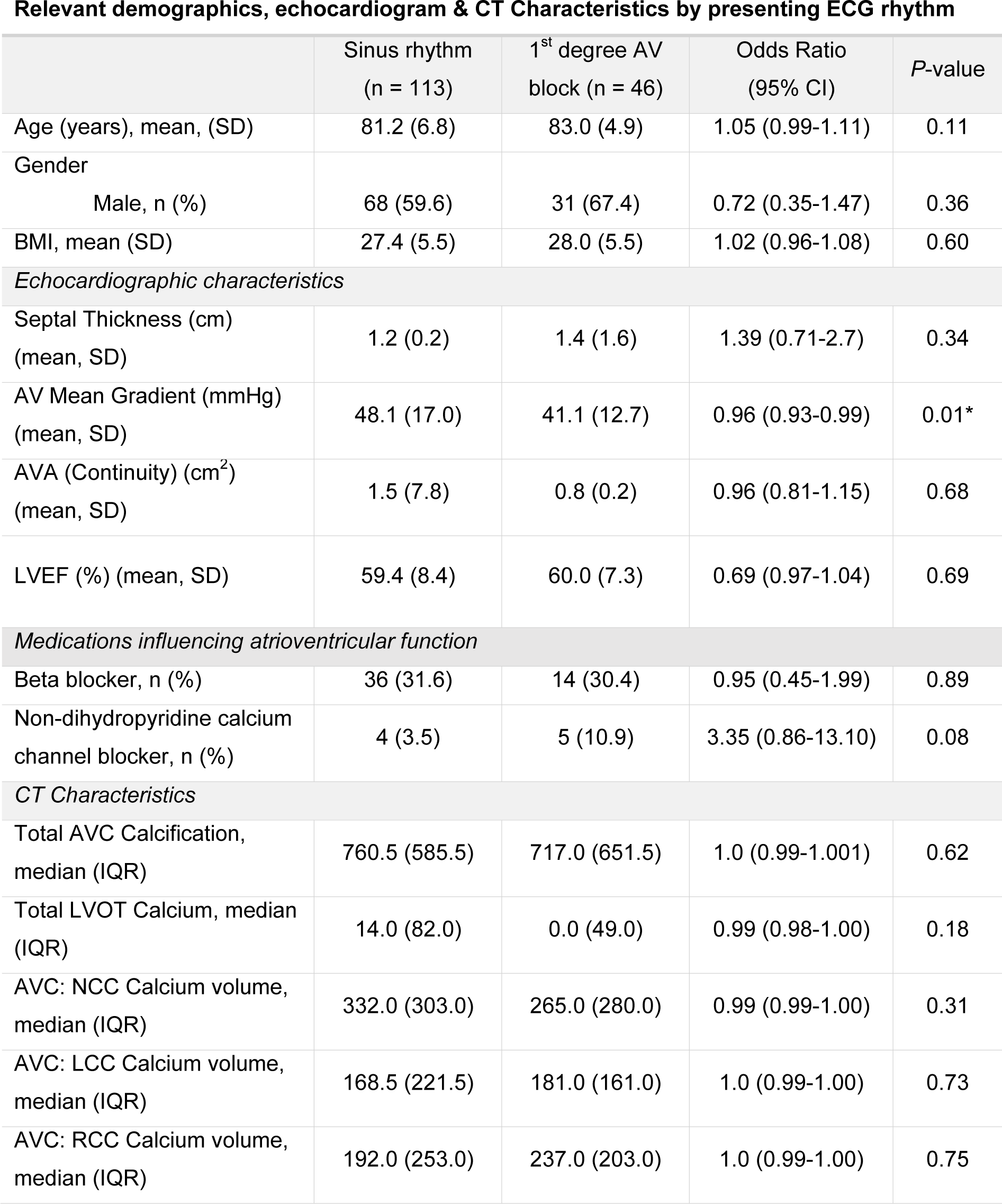
Relevant demographics and echocardiographic parameters by presenting rhythm. BMI: body mass index; AV: aortic valve; AVA: aortic valve area; LVEF: left ventricular ejection fraction; AVC: aortic valve calcium; SD: standard deviation; IQR: interquartile range. CT: computed tomography; AV: atrioventricular; STS: society of thoracic surgery; SD: standard deviation; CI: confidence interval

**Relevant demographics, echocardiogram & CT Characteristics by presenting ECG rhythm**
|  | Sinus rhythm<br>(n = 113) | 1<sup>st</sup> degree AV<br>block (n = 46) | Odds Ratio<br>(95% CI) | P-value |
| --- | --- | --- | --- | --- |
| Age (years), mean, (SD) | 81.2 (6.8) | 83.0 (4.9) | 1.05 (0.99-1.11) | 0.11 |
| Gender |  |  |  |  |
| Male, n (%) | 68 (59.6) | 31 (67.4) | 0.72 (0.35-1.47) | 0.36 |
| BMI, mean (SD) | 27.4 (5.5) | 28.0 (5.5) | 1.02 (0.96-1.08) | 0.60 |
| *Echocardiographic characteristics* |  |  |  |  |
| Septal Thickness (cm)<br>(mean, SD) | 1.2 (0.2) | 1.4 (1.6) | 1.39 (0.71-2.7) | 0.34 |
| AV Mean Gradient (mmHg)<br>(mean, SD) | 48.1 (17.0) | 41.1 (12.7) | 0.96 (0.93-0.99) | 0.01* |
| AVA (Continuity) (cm<sup>2</sup>)<br>(mean, SD) | 1.5 (7.8) | 0.8 (0.2) | 0.96 (0.81-1.15) | 0.68 |
| LVEF (%) (mean, SD) | 59.4 (8.4) | 60.0 (7.3) | 0.69 (0.97-1.04) | 0.69 |
| *Medications influencing atrioventricular function* |  |  |  |  |
| Beta blocker, n (%) | 36 (31.6) | 14 (30.4) | 0.95 (0.45-1.99) | 0.89 |
| Non-dihydropyridine calcium<br>channel blocker, n (%) | 4 (3.5) | 5 (10.9) | 3.35 (0.86-13.10) | 0.08 |
| *CT Characteristics* |  |  |  |  |
| Total AVC Calcification,<br>median (IQR) | 760.5 (585.5) | 717.0 (651.5) | 1.0 (0.99-1.001) | 0.62 |
| Total LVOT Calcium, median<br>(IQR) | 14.0 (82.0) | 0.0 (49.0) | 0.99 (0.98-1.00) | 0.18 |
| AVC: NCC Calcium volume,<br>median (IQR) | 332.0 (303.0) | 265.0 (280.0) | 0.99 (0.99-1.00) | 0.31 |
| AVC: LCC Calcium volume,<br>median (IQR) | 168.5 (221.5) | 181.0 (161.0) | 1.0 (0.99-1.00) | 0.73 |
| AVC: RCC Calcium volume,<br>median (IQR) | 192.0 (253.0) | 237.0 (203.0) | 1.0 (0.99-1.00) | 0.75 |

On multiple linear regression, there was no relationship between the primary outcome of total AVC calcium volume and PR, AH and HV intervals, when adjusted for age, gender and aortic valve mean gradient (Table 3).

**Table 3:** Multi-variable linear regression of the primary outcome (Total AVC Calcium volume) with markers of atrioventricular conduction (PR, AH and HV intervals) adjusting for age, gender and aortic valve mean gradient. EPS: electrophysiology study; AVC: aortic valve complex; LVOT: left ventricular outflow tract; NCC: non-coronary cusp; LCC: left coronary cusp; RCC: right coronary cusp

|  | Regression<br>coefficient (B) | Standard Error | P-Value |
| --- | --- | --- | --- |
| **Outcome: PR Interval** |  |  |  |
| Total AVC Calcium (mm<sup>3</sup>) | 0.006 | 0.008 | 0.476 |
| Age (years) | 0.505 | 0.462 | 0.277 |
| Gender | -7.107 | 6.661 | 0.288 |
| Aortic valve mean gradient (mmHg) | -0.286 | -0.136 | 0.138 |
| R<sup>2</sup>: 0.041 |  |  |  |
| F-Ratio: 1.501 (p = 0.205) |  |  |  |
| **Outcome: AH Interval** |  |  |  |
| Total AVC Calcium (mm<sup>3</sup>) | 0.006 | 0.008 | 0.410 |
| Age (years) | 1.273 | 0.467 | 0.007* |
| Gender | -7.071 | 6.700 | 0.293 |
| Aortic valve mean gradient (mmHg) | -0.244 | 0.193 | 0.293 |
| R<sup>2</sup>: 0.080 |  |  |  |
| F-Ratio: 2.981 (p=0.021) |  |  |  |
| **Outcome: HV Interval** |  |  |  |
| Total AVC Calcium (mm<sup>3</sup>) | 0.002 | 0.002 | 0.254 |
| Age (years) | 0.159 | 0.133 | 0.235 |
| Gender | -1.644 | 1.956 | 0.402 |
| Aortic valve mean gradient (mmHg) | -0.065 | 0.056 | 0.402 |
| R<sup>2</sup>: 0.033 |  |  |  |
| F-Ratio: 1.471 (p=0.213) |  |  |  |

\*Coefficients are unstandardized

### Mendelian randomisation study

In the primary IVW analysis, there was no evidence of a significant association between genetically predicted liability of AS and PR interval (beta: -0.41, 95% CI -1.33-0.51, p = 0.38; Figure 2). This was consistent on all sensitivity analyses including weighted median (beta: -0.08, 95% CI -0.57-0.42, p = 0.76), weighted mode (beta: -0.19, 95% CI -0.78-0.39, p = 0.53), and MR-Egger (beta: 0.86, 95% CI -1.52-3.24, p = 0.49). MR-Egger intercept was non-significant (Supplementary Table 1). There was significant heterogeneity identified by Cochran’s Q statistic (Q statistic: 124.92, p < 0.0001); nevertheless, similar neutral results were obtained after exclusion of outliers by both MR-PRESSO (beta: -0.24, 95% CI -0.80-0.33, p = 0.43) and Cook’s distance (IVW beta: -0.17, 95% CI -0.98-0.64, p = 0.68; Supplementary Table 2). Leave-one-out analyses are presented in Supplementary Figure 2, accompanied by scatter and funnel plots presented in Supplementary Figures 2 and 3, respectively. The more liberal SNP inclusion criteria of p < 5 x 10^-6^ led to an increase in the number of included SNPs from 16 to 56, yet the MR analysis results were similarly neutral (Supplementary Table 3). Further sensitivity analyses using a restricted subset of 5 SNPs that were additionally associated with aortic valve calcification based on computed tomography also yielded neutral results (Supplementary Table 5). The positive control analysis demonstrated expected results. A significant association was identified between genetically predicted liability of AS and heart failure (IVW odds ratio (OR) 1.16, 95% CI 1.09-1.24, p < 0.0001), with this positive result supported by all sensitivity analyses (Supplementary Table 4). On further evaluation of the relationship between genetically predicted AS and AV block, a significant association was identified on IVW analysis (OR 1.15, 95% CI 1.02-1.30, p=0.02), but this was not supported by the sensitivity analyses weighted-mode (OR 1.21, 95% CI 0.97-1.50, p=0.11), weighted-median (OR 1.15, 95% CI 1.00-1.33, p=0.052) nor MR-Egger (OR 1.13, 95% CI 0.79-1.61, p=0.52) (Supplementary Table 6).

**Figure 2:**
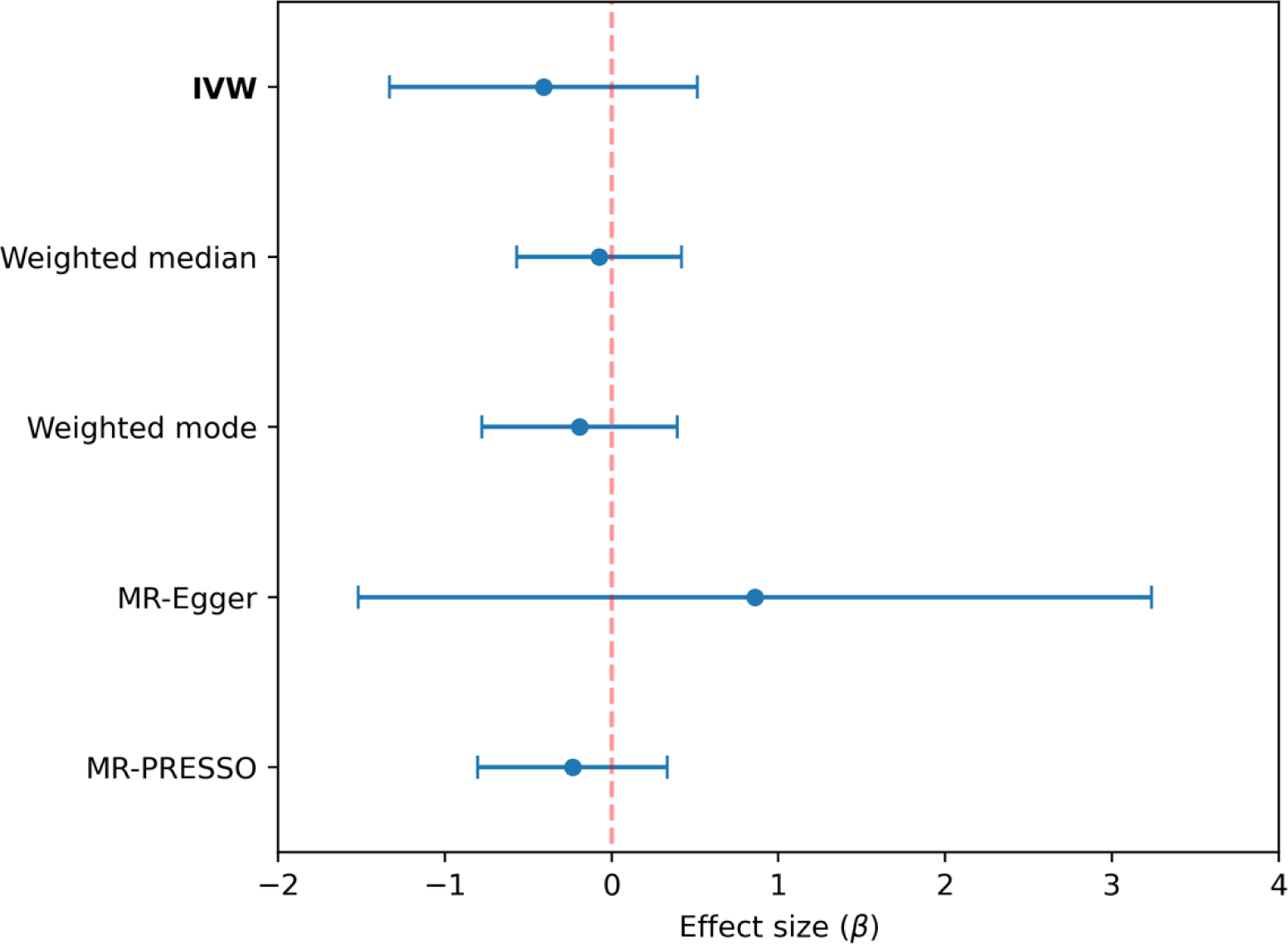
Mendelian randomization inverse variance weighted and sensitivity analysis estimates for the effect of genetically predicted liability of aortic stenosis on PR interval duration.

## Discussion

The present study used two separate approaches to evaluate the relationship between calcific aortic stenosis and AV conduction. The results demonstrate no evidence that the presence of AVC calcification is associated with prolongation of AV conduction. The cross-sectional observational study found no significant relationship between pre-defined calcium variables and first-degree heart block. Similarly, there were no clinically relevant associations between total AVC calcium volume and markers of atrioventricular conduction including PR, AH and HV duration. Concordantly on MR analyses, there was no significant association between genetically predicted liability for calcific AS and PR interval, and the association between genetically predicted AS and AV block was not robust to sensitivity analyses.

Despite a neutral result for the relationship between calcific AS and AV conduction disturbance, a substantial burden of bradyarrhythmia has previously been reported in patients with symptomatic severe AS. In a previous analysis of 40 patients with severe AS and syncope, AV block was the identified aetiology of syncope in 35%^16^, and in a prospective study of 106 severe AS patients investigated with one week of continuous ECG monitoring, 5.8% were identified to have HGAVB^17^, with similar findings in a study of 435 TAVI candidates undergoing 24-hour pre-procedural ECG monitoring^18^. One of the leading hypotheses for this has traditionally been disruption of the conduction system by fibrosis and calcium deposition. This hypothesis has been further supported by evidence that disruption of the AVC by TAVI^19^ and encroachment in the context of peri-annular complications of infective endocarditis^20^ similarly lead to conduction system disease.

Additionally some historical studies have supported the role of calcification specifically, including an analysis of 24 AS patients where 12 patients with aortic valve calcification had significantly longer HV intervals than 12 patients without calcification^4^. However other studies have demonstrated discordant findings. One analysis of 22 patients with syncope and AS found no association between aortic valve calcification and HV interval^3^. Similarly, Asmarats et al. showed no statistically significant association between increased Agatston calcium score and increased arrhythmic events in their study of 106 patients, albeit this analysis was not restricted to bradyarrhythmias^17^. The present study is a larger cohort that benefits from contemporary CT resolution with volumetric calcium characterisation and detailed invasive EPS data. Despite this, the current analysis similarly found no significant association between AVC calcium volume and PR, AH, or HV intervals. This was supported by the MR analyses which similarly demonstrated a neutral result for the relationship between genetically predicted liability for AS and PR interval, with concordant findings on sensitivity analyses including a restricted analysis using only SNPs which were associated with aortic valve calcification. While the association between genetically predicted AS liability and AV block was statistically significant on IVW analysis, the results of the weighted-mode, weighted-median and MR-Egger analyses were all non-significant and thus the IVW analysis should be interpreted with caution. Together, these results suggest that calcific AS may not have a causal role in AV conduction disease.

Similarly, the results did not support the hypothesis that asymmetric calcium deposition, particularly in the NCC and RCC regions, which are in closer anatomical proximity to the bundle of His, may predispose to prolonged AV node conduction due to localised infiltration. A possible explanation may be a fibrotic-predominant AS phenotype causing conduction disease, in lieu of calcium deposits. Although difficult to obtain, histopathological data would help better clarify the underlying mechanisms of this finding.

The main strength of the present study is the multimethodological approach, including a cohort of patients with prospectively characterisation of CT and EPS data, paired with an MR study involving several sensitivity analyses. Nevertheless, there are still several limitations to consider for both the prospective cohort and MR studies. In the observational arm, there was no control (non-AS) group available with dedicated CT and EPS data for comparison. Given that the MR study concordantly demonstrated an overall neutral result, it was an important complementary analysis to include a positive MR control, which expectedly demonstrated clear evidence for a robustly positive causal relationship between genetic liability for AS and heart failure, consistent with previous reports^21,22^.

It is important to note that all patients received sedation for the EP study which may have increased vagal tone, and approximately a third of patients used AV node blocking agents (beta blocker or non-dihydropyridine calcium channel blocker) at baseline. The PR and AH intervals are particularly prone to fluctuations in the autonomic nervous system, and it is possible that any potentially significant association was dampened by these medications. However, the HV interval is considered a more robust and reliable component of AV conduction and is also assessable in patients with atrial fibrillation, unlike the PR and AH interval. The present study could also be extended in future by inclusion of rapid and decremental atrial pacing (AV Wenckebach protocol) to further assess AV conduction by measuring the functional and effective refractory period.

There are additional limitations unique to the MR analysis. Specifically, the inclusion criteria for the AS exposure dataset were not strictly limited to severe AS, and so inclusion of mild and moderate AS cases may have diluted a true positive association. However, despite this the positive control analysis was noted to demonstrate a robust association between genetically predicted liability for AS and heart failure. Additionally, the primary MR analysis was not restricted to strictly calcific AS, since some proportion of cases may have been a non-calcific phenotype. The majority of patients with AS do demonstrate a calcific phenotype, but nevertheless this limitation was formally addressed by performing supplementary analyses using SNPs that, in addition to being significant for AS at the conventional threshold of p < 5 x 10^-8^, were also associated with valvular calcification at p < 0.05 as per the original analysis^11^. The present analysis used a two-sample MR approach with overlapping samples, most notably with overlap from UK Biobank and deCODE for the analysis between AS and PR interval. However the impact of this overlap is unlikely to be of practical significance for two reasons: first, only strong instrumental variables with F-statistic >10 were used, and it has previously been shown that overlapping samples is a more substantial issue in the context of weak instrument bias^23^; and secondly, bias due to sample overlap in two-sample MR increases the risk of false positive results, whereas the present study reports an overall neutral relationship.

## Conclusions

The present combined analyses did not support the hypothesis that AVC calcification increases the risk of AV nodal conduction disease. This result challenges the belief that local aortic valve calcification infiltrates and disrupts the conduction pathway in patients with AS. This is important in both shaping our understanding of conduction disease pathogenesis in the context of AS and should inform future work focused on predicting and reducing the risk of conduction disease after valvular intervention.

## Data availability statement

The data used in the Mendelian randomisation analyses are publicly available. Summary statistics for aortic stenosis are available from: https://doi.org/10.5281/zenodo.7505361; PR interval from: https://cvd.hugeamp.org; heart failure from: https://gwas.mrcieu.ac.uk/datasets/ebi-a-GCST90018806/; and AV block from https://r9.finngen.fi

## Acknowledgements

Nil.

